# Mendelian randomization linking metabolites with enzymes reveals known and novel pathway regulation and therapeutic avenues

**DOI:** 10.1101/2025.07.29.25332349

**Authors:** Adriaan van der Graaf, Sadegh Rizi, Chiara Auwerx, Zoltán Kutalik

## Abstract

Reactions between metabolites are catalyzed by enzymes. These biochemical reactions form complex metabolic networks, which are only partially characterized in humans and whose regulation remains poorly understood. Here, we assess human biochemical reactions and regulation using Mendelian randomization (MR), a genetic observational causal inference technique to understand the methods’ strengths and weaknesses in identifying metabolic reactions and regulation. We combine four metabolite and two protein quantitative trait locus (QTL) studies to determine how well MR recovers 945 curated canonical enzyme-substrate/product relationships. Using genetic variants from an enzyme’s transcribed (*cis*) region as instrumental variables, MR-inferred estimates have high precision (35%-47%) but low recall (3.2%-4.6%) to identify the substrates and products of an enzyme. Testing reverse causality from metabolites to enzymes using genome-wide instruments, yields lower precision (1.8%-8.5%) and recall (1.0%-1.9%) due to increased multiple testing burden. Literature review of 106 Bonferroni significant results identifies 45 links (43%) confirmed by different degrees of evidence, including bidirectional links between linoleate and Cytochrome P450 3A4 (CYP3A4) levels (P = 8.6 . 10^-32^). Eleven enzymes in the 106 links involve drug targets, allowing for an interpretation between N-acetyl putrescine and IL1RAP (P = 2.7 . 10^-7^), as IL1RAP is target of the psoriasis drug Spesolimab, and putrescine levels are elevated in psoriatic tissues. This work highlights how MR can be leveraged to explore human metabolic regulation and identify both canonical reactions and previously unknown regulation.

## Introduction

Metabolic reactions with unfavorable activation energy are catalyzed by enzymes. These enzymes are tightly regulated to maintain optimal metabolite concentrations under fluctuating environments, preventing both the depletion of essential metabolites and their accumulation to toxic levels. Enzymes can be regulated through processes such as competitive and uncompetitive inhibition, covalent enzyme modifications, phosphorylation and pathway inhibition or activation^1^. Yet, our knowledge of human metabolism remains incomplete: i) novel human metabolic pathways are still being discovered^2^, ii) the human genome still contains ‘orphan’ enzymes predicted to have a catalytic function that have yet to be experimentally demonstrated^3,4^ and iii) the regulation of the activation or repression of metabolic pathways is still an active field of study, for instance through metabolic flux analysis^5^.

It is possible to model metabolite pathways as a series of causal relationships, where the enzymatic activity of a protein is causal to the concentration(s) of the substrate and the product. This approach allows the identification of new putative metabolic links and regulatory mechanisms. These conclusions are usually derived from techniques such as randomized control experiments, metabolic flux analyses or Bayesian causal networks, and have successfully identified metabolic pathways in humans and other organisms^5–8^.

In this work, we explore a different causal inference technique to infer human pathway regulation: Mendelian randomization (MR). MR is an observational causal inference technique that uses genetic information to establish directional links between an exposure and an outcome^10,11^. In principle, it is possible to identify pathway regulation between metabolites and proteins using MR (Box 1). Motivated by the increased availability of well-powered metabolite (mQTL) and protein (pQTL) quantitative trait locus studies, as well as the ability of *cis* MR methodology to meaningfully improve drug target prediction^12,13^, we expect that MR is valuable in identifying the substrates and products of an enzyme and its regulation. To this end, we explore if and how MR can be used to derive meaningful conclusions in human metabolism and its regulation.

Here, we identify regulation between enzymes and metabolites through a form of observational causal inference called Mendelian randomization (MR). MR allows the identification of causal relationships between a heritable exposure and an outcome even when these traits are not measured in the same individuals by using genetic variants that influence an exposure of interest. Should there be a causal relationship between this exposure and an outcome, the genetics of the exposure should also be associated to the outcome in a proportional manner. MR operates on three main assumptions: i) *The relevance assumption*: the genetic variant needs to be robustly associated to the exposure of interest. ii) *The independence assumption*: the genetic variant needs to be independent from any confounder of the exposure-outcome relationship and iii) *The exclusion restriction:* the genetic variant needs to affect the outcome only through the exposure, and there should be no other paths. Violations of the exclusion restriction assumption – otherwise known as horizontal pleiotropy – is difficult to account for, especially when there is only a limited number of independent genetic variants available^11,14^. Recent advances in MR allow for pleiotropy-robust analysis of molecular phenotypes that have a single or only a handful of associated regions^15,16^.

Our goal in this study is to identify to which extent enzymatic reactions can be reidentified by MR based on available pQTL and mQTL studies. Then, we aim to study enzyme-metabolite pairs significantly linked by MR but are not in the gold standard benchmark set to estimate if these reflect true human biology or rather false positives. For this, we use two pathway references to build a metabolic map between metabolites and their enzymes in humans (**Figure 1**). Then, using pQTLs derived from two independent studies ^17,18^ and mQTLs derived from four independent studies ^19–22^ (**Figure 1a**), we estimate the causal effects that proteins have on metabolites and vice versa, using the protein levels as a proxy for enzymatic activity, distinguishing between MR results based on instruments in the *cis* region of the enzyme and those based on meta-analyzing all *cis* and *trans* regions together (**Figure 1b**). We find that MR methods have limited discriminative ability to determine if a metabolite is catalyzed by a specific protein. Yet, Bonferroni-significant MR estimates are enriched for true reactions. Extensive literature search of the Bonferroni significant combinations that are not in our metabolic map proposes possible mechanistic explanations for some of the newly identified links and points out multiple drug targets among the involved enzymes, suggesting putative metabolite downstream drug effects.

**Figure 1.**
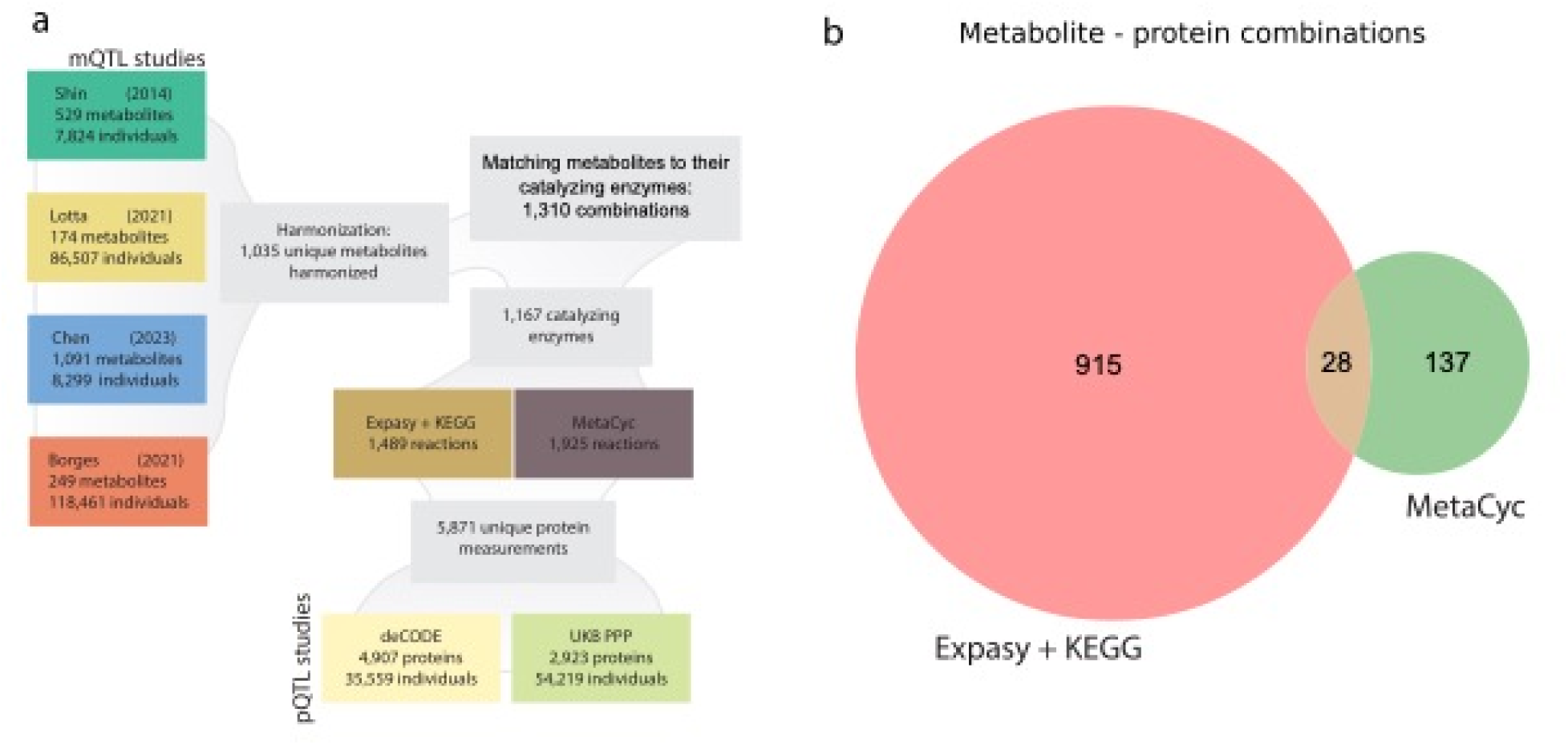
Overview of the data used in this study. (a) The different metabolite quantitative trait locus (mQTL) studies and protein quantitative trait locus (pQTL) studies, combined with two pathway reaction references ‘Expasy + KEGG’ and ‘MetaCyc’. Based on the matching of proteins to metabolites we identify 1,742 enzyme metabolite links. (b) The number of measured enzyme metabolite links present in each pathway reference.x

## Methods

### Metabolite quantitative trait locus studies

We analyzed four metabolite quantitative trait studies, as described in our previous publication^16^. In short, the summary statistics for four mQTL studies were downloaded from their respective resources^19–22^. and the metabolites were harmonized to Human metabolite database (HMDB) identifiers. This resulted in 1,109 metabolites to which an HMDB ID was assigned.

### Protein quantitative trait locus studies

We analyzed two pQTL studies: the DeCode study^18^ and the UK biobank Pharma Proteomics Project (PPP)^17^. The DeCODE study comprises genetic associations of 4,907 plasma proteins across 35,559 individuals. The PPP study contains genetic associations for 2,923 plasma proteins across 54,219 individuals. The summary statistics and meta data were downloaded from their respective resources, and were harmonized using the UniProt identifiers, which were provided by all three studies.

### Identification of catalytic enzymes

We used two independent resources to build our metabolic map: KEGG+Expasy and Metacyc. For KEGG+Expasy, we used the Expasy enzyme resource to retrieve the enzymes that have catalytic effects in humans^23^, and the KEGG resource to identify metabolites that are involved in the reactions. Out of 8,227 enzymes from the Expasy resource, we identified 4,277 enzyme commission (EC) numbers that match to human enzymes, including 3,592 unique enzymes (some enzymes catalyze multiple reactions)^24^. We used KEGG to match the compounds to each enzymatic reaction. We identified 229 human KEGG pathways in which the measured compounds are present and downloaded the XML files for each human pathway. From these, we derived the 1,489 reactions for which protein and reactant information was available for at least one pQTL and mQTL study. After matching these reactions to the enzymes that catalyze them, this resulted in 1,236 measured enzyme-reactant combinations. As a secondary metabolite reference, we used HumanCyc v24.0 database to identify enzymes and reactants that are present in a pathway ^25^. Matching a total of 1,925 unique reactions that can be catalyzed by 2,933 unique proteins. After matching the reactions to the measured proteins and metabolites we ended up with 231 unique exposure and outcome combinations.

### Summary statistics harmonization

We jointly harmonized summary statistics from all pQTL and mQTL studies in the same way: First, if necessary, the files were lifted over to human genome build GRCh37 to match our allele reference using the UCSC liftover tool (https://genome.ucsc.edu/cgi-bin/hgLiftOver). Second, variants were matched to the UK10K linkage disequilibrium reference panel^26^, matching variants on their genomic positions and alleles, while removing palindromic variants and variants that have a minor allele frequency less than 0.5% in the panel.

We then harmonize the effect sizes of the summary statistics to the standard deviation per standard deviation scale:

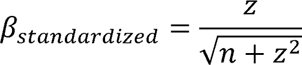

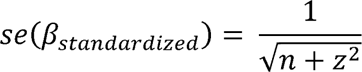

Where *n* is the variants sample size and *z* the Z score of the association. We set *n* to be the maximum sample size if *n* was not available for a given variant. The P value and effect size sign of the variant-trait association was converted into a Z score if Z score was not available^16^. If per variant sample size was available, we removed variants that were missing more than 5% of the maximum sample size in a region.

### Identification of associated regions

We identified associated regions by P value clumping genetic variants at a P value threshold of smaller than 5. 10^-8^, using the plink (v1.90b7)^27^ –clump command with a clumping window of 250 Kb and an LD threshold of 0.01 r^2^. Overlapping clumped regions were combined. Non-overlapping regions were analyzed by all four *cis* MR methods^16^. If a method requires instrumental variables, these were clumped from the full region using the same P value and LD threshold. To reduce the dependence of reverse causality, we removed the protein *cis* region, if the protein is the outcome in the meta-analysis.

### MR methods used in this study

We used four MR methods: MR-IVW^11^, MR-IVW-LD, MR-PCA^28^ and MR-link-2^16^. These methods have been chosen because they are appropriate to use on one or multiple associated regions. Code implementations for all the methods can be found at (https://github.com/adriaan-vd-graaf/mrlink2).

We performed three ways of interpreting the MR analyses here: i) *cis*-MR, ii) *cis* + *trans* MR, and iii) meta-analyzed bidirectional meta-analysed MR: performing region-based MR for every exposure associated region in both protein-to-metabolite and metabolite-to-protein directions and meta-analyzing the region-based MR estimates.

In *cis* MR, we performed regional MR when any clumped variants in the gene region (defined in the section *Identification of associated regions*) is within 250Kb of the start or end of the gene that transcribes the enzyme according to Ensembl biomart (https://www.ensembl.org/info/data/biomart/index.html).

In the *cis* + *trans* analysis, we analyze all associated regions of an enzyme and treat each individual associated region as an independent MR analysis. Here we only test the direction from the enzyme to the metabolite.

In the meta-analyzed MR we perform a bidirectional MR identifying causality between enzymes and metabolites, and between metabolites and enzymes. This can identify canonical metabolic reactions (from the enzyme onto the metabolite), but also their regulation (both the enzyme onto the metabolite and the metabolite onto the enzyme). We perform an MR for all genetic associations of an enzyme or a metabolite and we meta-analyze these estimates. The enzyme to metabolite direction is the only direction that we consider truly causative, but as we perform biological follow up in both directions, we include both directions in the final approach.

### Discriminative ability analysis using AUC

We used the AUC to assess the discriminative ability of different MR methods. True links were defined as enzymes and their reactants, whereas the true non-links are all the other protein-metabolite combinations. The AUC was calculated with the scikitlearn library ^29^.

### Manual literature search and classification of evidence

For all MR-link-2-significant link we performed a literature review. Specifically, we searched for both the protein-metabolite combination and the metabolite in combination with the products and substrates of the reaction that the protein catalyzes in PUBMED, OMIM and the BRENDA enzyme database^30,31^. We classified our findings into five categories of evidence:

1. Canonical reaction: significant combinations where the exposure is the enzyme that directly catalyzes the metabolite outcome.
2. Reverse canonical evidence: causal relationships where the metabolite acts on their catalyzing enzyme.
3. Pathway evidence: causal relationships where the metabolite and the enzyme are in the same pathway.
4. Homologous evidence: causal relationships where a paralog (similar gene in the same organism) or ortholog (similar gene in another organism) is related to the metabolite. Which indicates that there is a strong effect of based on the BRENDA data resource).
5. Supporting evidence: These combinations are cryptic and may represent novel and unknown regulation, it is however also possible that these combinations are false positives.

### OpenTargets matching drugs to their consequences

We downloaded the ‘molecule’ and ‘diseases’ tables from OpenTargets platform (https://platform.opentargets.org/downloads; June 2024) and matched 1,550 OpenTargets drug targets to 358 of the unique enzyme measurements. We conducted a literature search of all drug targets that are Bonferroni significant in our original analysis.

## Results

### Matching pathway references with protein and metabolite QTLs

To build a ground truth dataset of enzymatic reactions that can be matched to in pQTL and mQTL studies, we first use two human pathway reference datasets to identify enzymes and their reactants in our metabolic map: KEGG+Expasy and MetaCyc to build a metabolic map^25,32,33^ (**Methods**). KEGG+Expasy contains 1,489 reactions, while MetaCyc contains 1,925 reactions (**Figure 1a**). Each reaction consists of one enzyme, one or more substrates, and one or more products (**Supplementary Table 1**) (**Methods**). We matched enzymes from our metabolic map to two protein QTL studies: the DeCODE study comprising 4,907 proteins across 35,559 individuals^18^ and the UK biobank Pharma Proteomics Project (UKB-PPP) comprising 2,923 proteins across 54,219 individuals (**Figure 1a**). After matching proteins to our metabolic map, 2,696 and 2,180 protein measurements had catalytic activity according to KEGG+Expasy and MetaCyc, respectively (**Figure 1a**). Across pQTL studies, 1,624 unique proteins had a catalytic function according to at least one pathway reference (**Supplementary Table 2**). We matched the metabolites from our metabolic map to those in four mQTL studies: Shin *et al.,* Lotta *et al.,* Chen *et al.*, and Borgess *et al.*^19–22^. This resulted in 324 unique enzymes across 424 measurements in reactions with 121 unique metabolites across 214 measurements. Overall, our metabolic map encompasses 1,310 protein-metabolite pairs, including 540 and 770 pairs in which the metabolite acts as substrate and product, respectively (**Figure 1a**) (**Figure 1b**) (**Supplementary Table 3**).

When considering these ground truth reactions from the metabolic map, enzyme abundances to reactant concentration are the true positives, as we expect that enzyme abundance causally influences the concentrations of substrates and products. We make a distinction between the enzyme-onto-metabolite direction versus the metabolite-onto-enzyme direction as the enzyme-onto-metabolite direction can be understood as directly causative: An enzyme converts a metabolite in a well understood physical process. As a reaction occurs, we expect substrate depletion and an increase in product concentration. Under the assumption that enzymes catalyze a reaction in a single direction, we expect to see negative causal estimates from the enzyme to the substrate and positive causal estimates from the enzyme to the product. In contrast, we do not consider the metabolite-onto-enzyme direction directly causative as there is no obvious physical process through which the reactant influences the abundance of the catalyzing enzyme. Instead, we consider such influences as indirect regulatory effects, such as for instance the regulation between cholesterol concentrations and the degradation of HMG-CoA reductase^34^, as well as between xenobiotics and cytochrome P450 3A4 abundances^35^.

### Mendelian randomization analyses

We perform MR using four *cis* MR methods (MR-IVW, MR-PCA, MR-IVW-LD and MR-link-2) to test how well MR can identify the causal relationships between an enzyme and its substrates and products (**Methods**). We consider three forms of MR analysis: i) *cis*-MR, where only the genetic variants proximal to the gene encoding for the enzyme of interest are used as instrumental variables in the MR; ii) *cis*+*trans* MR, which performs MR independently for each associated locus; and iii) bidirectional *cis*+*trans* meta-MR which meta-analyzes region-based MR estimates (**Methods**).

First, for the *cis*-MR analysis, we identify enzyme-to-metabolite effects using only the *cis* region (where the enzyme is transcribed) as a source of MR instruments Secondly, In *cis+trans* MR, we perform MR in each enzyme-associated region separately yielding multiple (region-specific) causal effect estimates. These MR effect estimates are tested against the gold standard set independently.

Thirdly, for meta-analyzed MR, we combine the individual region-based MR estimates from the *cis+trans* MR for each enzyme-to-metabolite MR, as well as for each metabolite-to-enzyme MR using inverse variance weighting meta-analysis (**Methods**).

We only test the enzyme to metabolite direction when performing *cis* and *cis*+*trans* MR, as we are testing the merit of these methods for identifying enzymatic reactions and a *cis* region is not available for a metabolite. In contrast to the meta-analyzed MR, where we also perform the metabolite-to-protein direction, as here we broaden the research question, and assess any potential indirect regulation through literature search.

In all these estimates, we consider the protein to metabolite direction to be the only truly causative direction, and we perform literature follow up on significant findings only for the most conservative, meta-analysis MR approach.

#### Cis Mendelian randomization to identify reactants and their enzymes

We applied MR to *cis* regions that associate (SNP P ≤ 5 . 10^-8^) to 159 enzymes (197 measurements) that are in the metabolic map. These enzymes catalyze reactions involving 88 metabolites (139 measurements), leading to 499 protein-metabolite pairs that are the positive controls in this study (**Supplementary Table 4**). All other 25,420 possible pairwise comparisons between enzymes and metabolites are considered as negative controls. We provide an example *cis* MR result in **Figure 2**. Here, arginase 1 converts arginine into ornithine and urea (EC 3.5.3.1) (**Figure 2a**) along with the MR-link-2 results, where the product causal direction is (**Figure 2b-d**).

**Figure 2.**
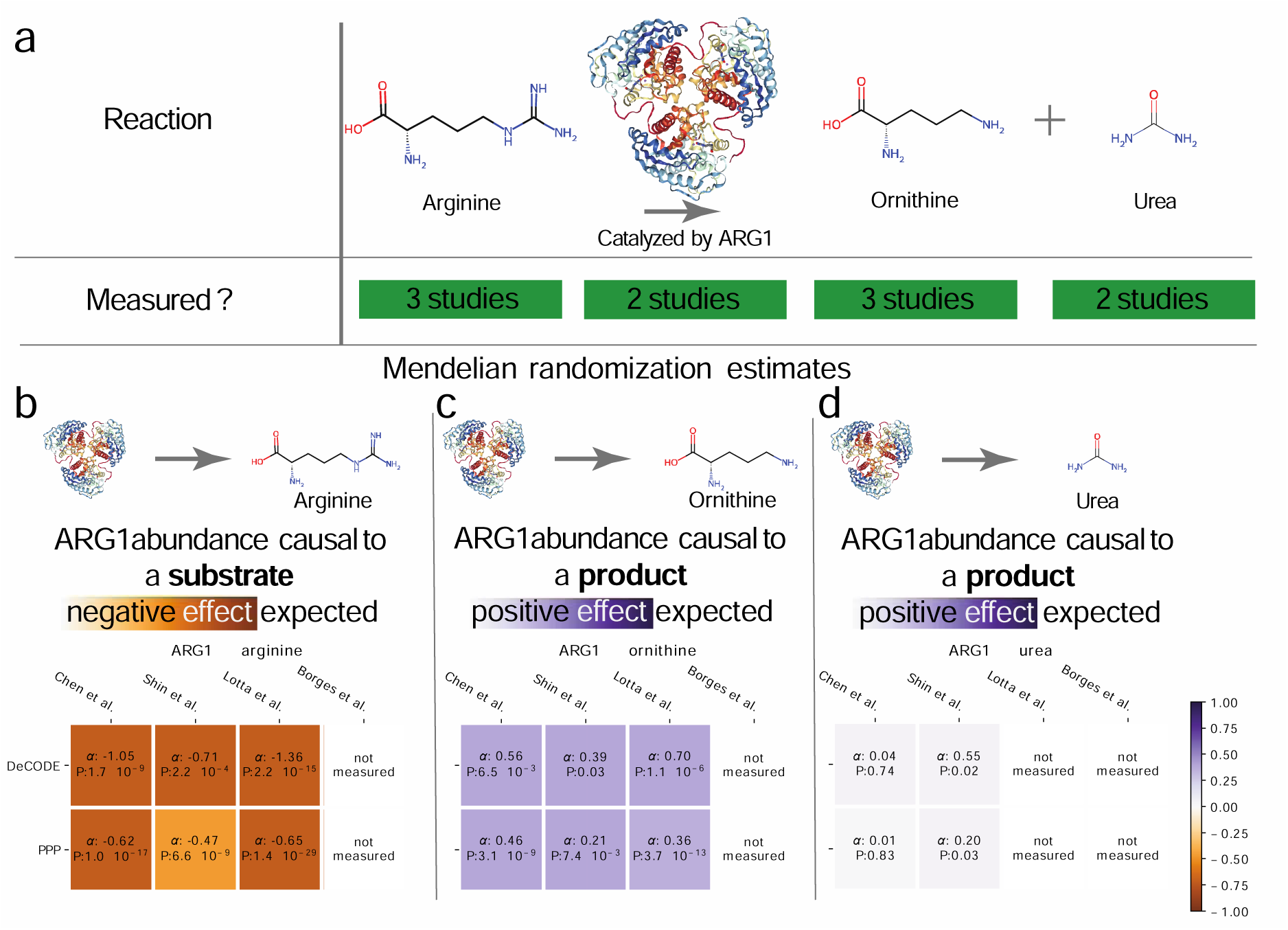
Example of MR effect estimation. (a) Considering the reaction catalyzed by arginase 1 (ARG1), with arginine as a substrate, and ornithine and urea as the product, one can estimate a causal relationship between the enzyme and the reactants, provided they are measured in each study. (b) The causal estimates between ARG1 and the arginine substrate, as arginine is a substrate a negative causal effect is expected. (c) The causal estimates between ARG1 and the reaction product ornithine. (d) The causal estimates between ARG1 and the reaction product urea. It is not possible to estimate a causal effect if either the enzyme or the metabolite is unmeasured in a single study

As *cis* regions are only available for enzymes, this methodology will only be able to identify causality from enzyme concentrations onto metabolite levels. When testing for the directionality of MR methods on substrates and proteins. MR-link-2 is the only method that identifies a significantly larger causal effect for products than for substrates (Kruskal-Wallis P = 0.011) (median product causal effect estimate α̂: 0.008, median substrate α̂: = -0.013) across 219 substrates and 320 products from our metabolic map (Supplementary Figure 1).

Using the positive control enzyme-metabolite combinations, it is possible to determine the discriminative ability of different MR methods, which is limited, with area under the receiver operator characteristic curve (AUC) ranging between 0.543-0.554 (**Supplementary Table 5**) (**Supplementary Figure 2**). We found that precision (proportion true positives over all detected) for all MR methods was highest in the low P value range, we show precision and recall (proportion of true positives over all positives) on a log scale (**Supplementary Figure 3a**) (**Supplementary Figure 3b**). Analyzing causal estimates below the *cis-*specific Bonferroni threshold 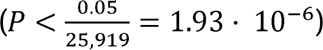, all methods had low recall 3.2%-4.6% combined with high precision 35%-47% (**Supplementary Figure 3**). Each MR method had similar precision and recall at any specific P value rank, with MR-IVW-LD having the highest recall (with lower precision) and MR-link-2 the highest precision (with lower recall), in line with our previous work^16^. Considering only unique enzyme-to-metabolite combinations, MR-PCA identified the largest number (n = 28) of unique enzyme-to-metabolite links, including 12 positive controls (**Supplementary Figure 3c**), and 16 likely false positives (**Figure 3d**). In contrast, MR-link-2 identified eight positive controls (**Figure 3c**) and five likely false positives (**Figure 3d**) (**Supplementary Figure 3**).

**Figure 3.**
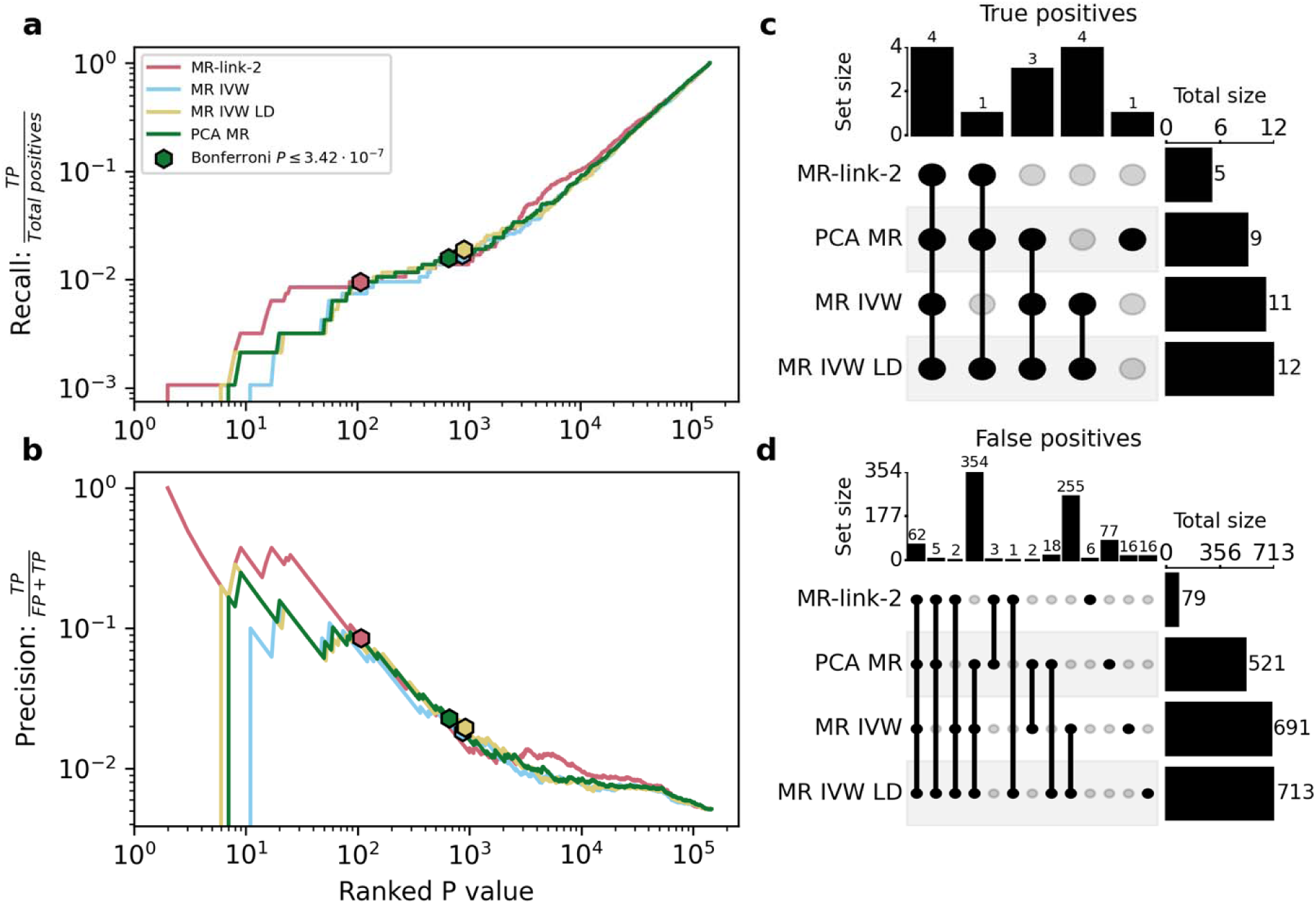
Precision and recall curves of MR methods when meta-analyzing results together in a bidirectional manner between enzymes and metabolites. (a) Recall and (b) precision depending on the chosen P value threshold ordering. (c) Set memberships for the true positives at each methods’ Bonferroni threshold. (d) Set memberships of false positives at each methods’ Bonferroni threshold. Abbreviations: (TP) True positive, (FP) False positive.

To extend the true positive set, we added further links that were reported in the BRENDA database^31^ (**Methods**). We determined for each enzyme-metabolite combination whether any of the metabolites had been found as a substrate, product or inhibitor in the enzymes that were otherwise missed by our original true positive dataset. Two out of five unconfirmed links found by MR-link-2 were an inhibitor or a substrate according to BRENDA. In the case of MR-PCA, 4 out of 16 links represented a substrate, product or inhibitor relationship.(**Supplementary Table 6**) (**Supplementary Figure 4**) (**Methods**)^31^.

### *cis* + *trans* MR to extend causality

Considering that the *cis* MR results had high precision with relatively low recall, we considered that including *trans* associated loci could increase the number of causal links that may be biologically informative. We set out to investigate the ability for *cis* MR methods to identify further protein-to-metabolite causality using all associated regions (*cis* + *trans* )

After analyzing a total of 451,502 combinations, of which 4,987 are considered true positives. At a Bonferroni significance threshold 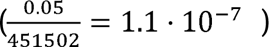 recall ranged between 0.4% and 1% depending on the MR method (**Supplementary Figure 5a**), while precision ranged between 11.6% and 1.8% (**Supplementary Figure 5b**) (**Supplementary Table 7**). Even though we expected to increase recall in the *trans* analysis over the *cis* analysis, at the cost of precision, the inclusion of separate trans region-based MR estimates identified fewer unique true positives for MR-link-2 the methods compared to the *cis* MR analysis. This is likely due to: the increased multiple testing burden, and an increase in the set of positives, as not all positives can be tested in the *cis* analysis. For illustration, MR-link-2 identified eight unique true positives in the *cis* analysis, compared to seven in the *cis*+*trans* analysis (**Supplementary Figure 3c**) (**Supplementary Figure 5c-d**) (**Supplementary Table 7**). Nonetheless, including *trans* regions in an analysis is not without merit: Some *trans* regions were individually significant at the stringent multiple testing threshold: for instance, ARG1 abundances were regulated in *trans* on chromosome 19, providing a concordant Bonferroni significant true positive causal estimate on ornithine concentrations (α̂ = 0.67, P = 4.0 . 10^-8^) (**Supplementary Table 7**).

### Meta-analyzing MR between enzymes and metabolites

Based on the information that *cis* as well as *trans* regions can be informative, we decided to combine all associated regions into a single causal estimate per enzyme-to-metabolite pair, as well as in the reverse direction: additionally testing the effect of metabolites on enzyme abundances using the standard inverse variance weighting meta-analysis (**Methods**). This is the first analysis where we include the reverse metabolite-to-enzyme regulation direction. we still only consider the enzyme-to-metabolite direction causative and attribute any significant results in the metabolite-to-enzyme direction to indirect regulation, which we later follow up in literature search. Combining all enzyme-metabolite pairs resulted in 215,556 combinations (506 enzymes * 213 metabolites * 2 directions), out of which 146,062 could be instrumented by MR, setting our meta-analysis Bonferroni-corrected threshold for significance at (P < 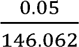 = 3.42 . 10^-7^) (**Methods**).

Of these, 945 enzyme-to-metabolite combinations represent ground truth positive controls in our metabolic map (**Supplementary Table 8**). We find that all tested MR methods in meta-analysis have modest discriminative ability (AUC: 0.512-0.531) (**Supplementary Table 9**). When considering Bonferroni-significant results, there is strong enrichment for true metabolite reactions across all methods (odds ratio: 2.9-13.2, P: 3.0 . 10^-4^- 7.4 . 10^-8^) (**Supplementary Table 10**). The differences in precision and recall between MR methods was more pronounced compared to *cis*-MR (**Figure 3a**) (**Figure 3b**); MR-link-2, MR-PCA, MR-IVW, and MR-IVW-LD identified 106, 660, 870, and 912 Bonferroni-significant effects, respectively. Methods differ strongly in their precision (**Figure 3a**) (**Supplementary Table 8**), with the best precision obtained by MR-link-2 (8.5%) compared to up to 2.3% for other methods. This comes at the cost of lower recall: nine out of 945 true comparisons pass Bonferroni significance with MR-link-2, compared to 19 for MR-IVW-LD (**Figure 3b**) (**Supplementary Table 8**).

These precision and recall curves can also be used to compare MR methods. Generally, all methods have similar precision and recall, but there are deviations, especially when considering the most significant parts of the curves. (**Figure 3a**) (**Figure 3b**) (**Supplementary Table 8**)

### Significant meta-analyzed MR-link-2 results show different types of evidence for pathway regulation

Since our ground truth dataset of reactions is not representative of the full biology between the metabolites and enzymes, we performed a literature review to annotate an MR-derived positive set of candidate reactions that were not reported in our metabolic map. We expected meta-analyzing region-based MR estimates be the most robust analysis method and MR-link-2 yielded the highest precision, we manually annotated all 106 (across 84 unique combinations) MR-link-2 Bonferroni-significant meta-analyzed results (**Figure 4c,d**) (**Figure 5**). We classified identified links into five classes: i) “evidence of a canonical reaction” (n = 12, 11%), ii) “reverse canonical evidence” (i.e., the metabolite influences the enzyme, n = 8, 7.5%), iii) “pathway evidence” (i.e., the metabolite and enzyme share a pathway, n = 6, 5.6%), iv) “homologous reaction evidence” (i.e., the enzyme was shown to interact with the metabolite based on homologous enzymes, n = 4, 3.8%) and v) “supporting evidence” (i.e., biological evidence present that the enzyme and metabolite are related in model organisms or shared biological pathways, n = 15, 14%). The manual annotation of the Bonferroni significant results excluding “supporting evidence” are shown in **Table 1**, and all annotations are visualized as a graph in **Figure 4**. The manual annotations of the significant MR-link-2 meta-analyzed results that include “supporting evidence” are listed in **Supplementary Table 11**.

**Figure 4.**
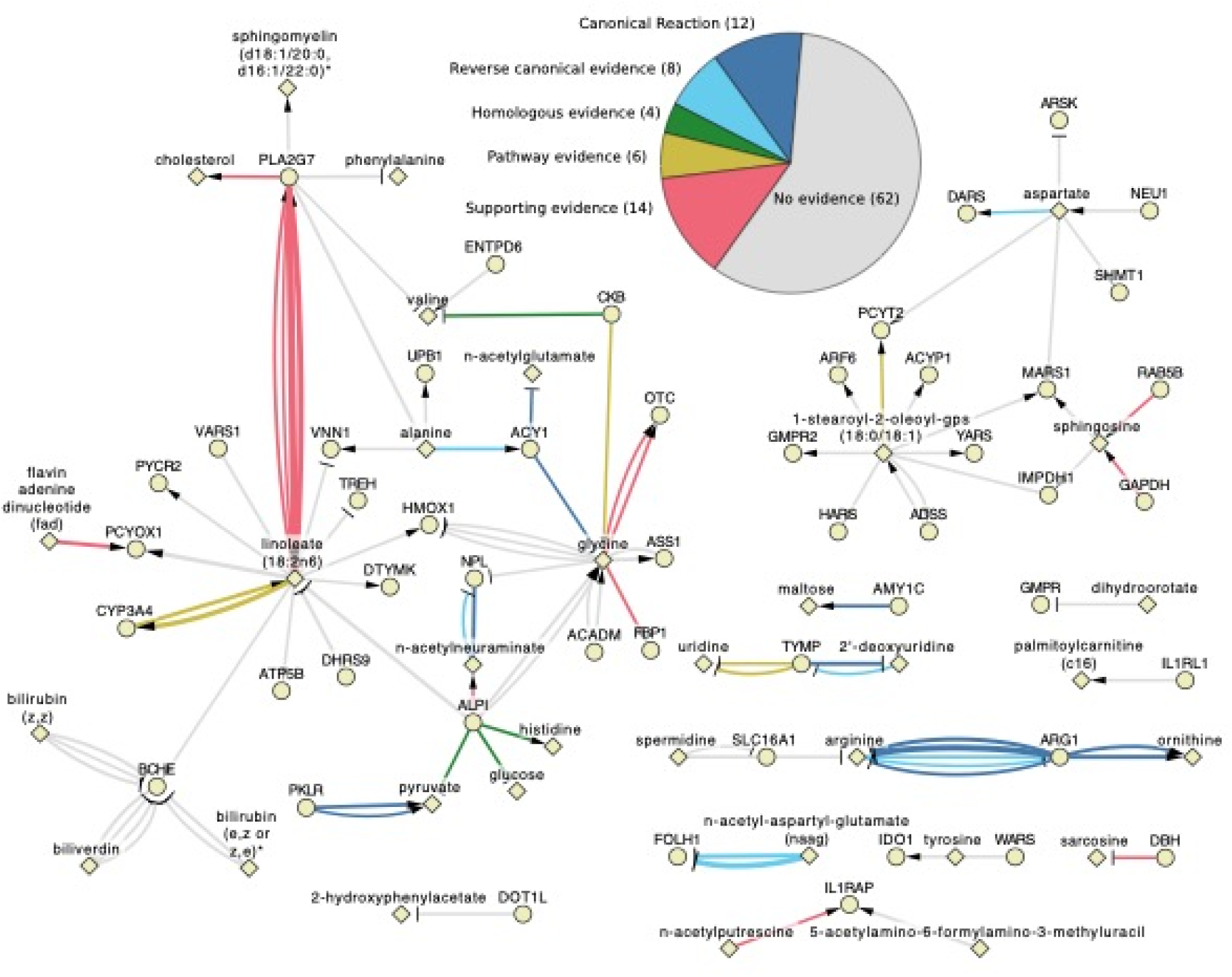
Bonferroni significant MR-link-2 results with their biological. The pie chart depicts the numbers found for each class of interpretation and serves as a legend. The metabolites are depicted as diamonds, while proteins are depicted as circles. A blunted arrow indicates a negative causal relationship, while a pointy arrow indicates a positive causal relationship.

**Table 1:**
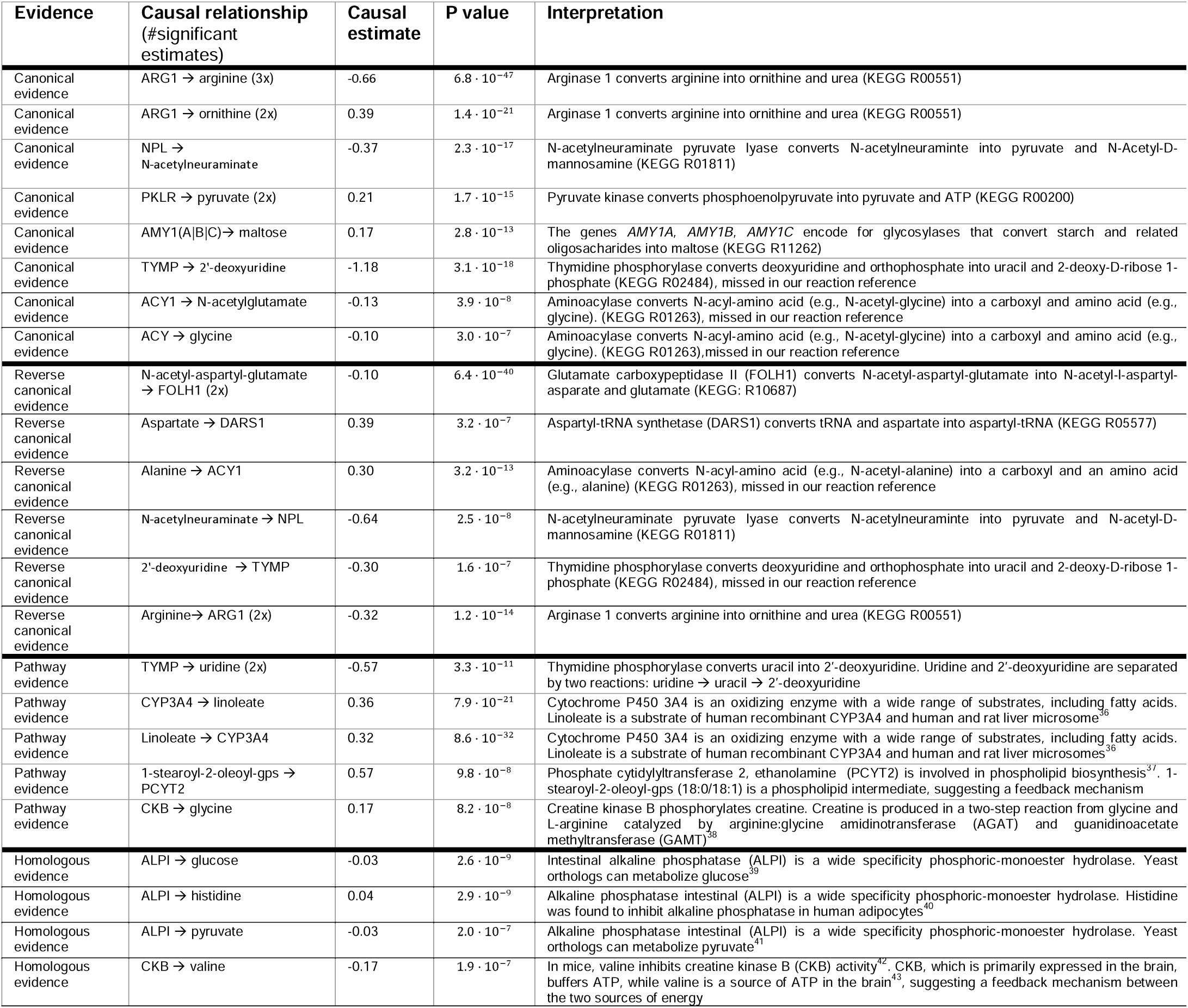
Bonferroni significant MR-link-2 results with supporting evidence. Causal relationships are classified according to whether there is evidence of a canonical reaction, reverse canonical evidence (the metabolite influences the enzyme), pathway evidence (the metabolite and enzyme share a pathway), homologous reaction evidence (the enzyme has been shown to interact with the metabolite from homologous evidence). Enzymes or metabolites might be measured in multiple studies, leading to multiple estimates (indicated by a ‘2x’ or ‘3x’ in the “Causal relationship” column). We only report the most significant result and the number of Bonferroni-signifcant (P < 3.4 · 10^-7^) combinations. Full results in Supplementary Table 11.

#### Canonical reactions

Twelve Bonferroni-significant protein-to-metabolite effects represent canonical reactions where the enzymes are estimated to causally influence the substrate or the product of the reaction (**Table 1; Figure 5**). Three of these direct reactions were missed by our metabolic map due to differences in harmonization of compounds and reactions and retrieved through manual curation: TYMP (thymidine phosphorylase) causal to 2’- deoxyuridine levels (α̂ = -1.2, P = 3.1 . 10^-18^), ACY1 (aminoacylase 1) causal to N-acetyl-glutamate levels (α̂ = -0.13, P = 3.9 . 10^-8^) and ACY1 causal to glycine levels (α̂ = -0.13, P = 3.9 . 10^-8^).

#### Reverse Canonical Reactions

MR-link-2 identifies eight metabolites that influence their metabolizing enzyme (**Table 1**) (**Figure 5**) (**Supplementary Table 11**). These reactions can be further divided depending on whether the enzyme also causally influences the metabolite (i.e., bidirectional effects; n = 4; arginine ➔ ARG1 (2x), N-acetyl-neuraminate ➔ NPL and 2’-deoxyuridine ➔ TYMP (**Table 1**)), or not (i.e., unidirectional effects; n = 4; (2x) N-acetyl-aspartyl-glutamate (NAAG) ➔ FOLH1, aspartate ➔ DARS and alanine ➔ ACY1). In the latter scenario, all tested enzymes had an instrumental variable, and the MR estimate from enzyme to metabolite was non-significant (**Supplementary Table 8**). This suggests existence of feedback mechanisms wherein metabolites sense and regulate enzyme levels. In such examples, NAAG has a negative effect on its catalyzing enzyme, folate hydrolase (FOLH1, also known as glutamate carboxypeptidase 2; measured in two studies: α̂ = -0.10, P = 6.4 . 10^-40^ and α̂ = -0.07, P = 1.6 . 10^-25^). The enzyme converts NAAG into N-acetylaspartate and glutamate, suggesting a potential feedback mechanisms that would prevent accumulation of glutamate, which has neurotoxic properties at high concentrations^44,45^ (**Table 1**).

#### Pathway regulation

We identify five enzyme-metabolite pairs that are closely related in the same pathway but not connected through a canonical reaction (**Table 1**) (**Figure 5**). For instance, TYMP ➔ uridine (uridine was measured twice, two causal estimates: α̂ = -0.57, P = 3.2 . 10^-11^ and α̂ = -0.55, P = 5.6 . 10^-8^), is two reactions away from 2’-deoxyridine, a TYMP substrate (uridine ➔ uracil ➔ 2’-deoxyuridine) (KEGG: hsa01232)^32,33^. We also found that creatine kinase B (CKB) concentration causally affects glycine levels (α̂ = 0.16, P = 8.2 . 10^-8^). Creatine is a substrate of CKB and is two reactions away from glycine (glycine ➔ guanidinoacetate ➔ creatine)^38^. Interestingly, no MR method identified a causal relationship between CKB and creatine ( P >0.09) (the reverse direction) (**Supplementary Table 8**), suggesting that the causal link between glycine and creatine may represent a regulatory mechanism.

#### Homologous evidence

Sometimes, a homologous enzyme has activity that can be related to the metabolite that MR-link-2 implicates. This can either be through activity in another organism (orthologous enzyme) or through activity in a copy of the gene (paralogous enzyme). We identify that alkaline phosphatase, intestinal (ALPI) has a causal effect on three metabolites: glucose (α̂ = -0.03, P = 2.6 . 10^-9^), pyruvate (α̂ = -0.03, P = 2.0 . 10^-7^) and histidine (α̂ = 0.04, P = 2.9 . 10^-9^). ALPI is a wide-acting phosphatase, and studies of the yeast ortholog have shown that pyruvate and glucose can be a reactant of the enzyme^39,40^ (**Table 1**). In line with this, ALPI levels have been found as risk factors for diabetes^46,47^. We also estimated that CKB causally lowers valine concentration (α̂ : -0.187 P: 1.9 . 10^-7^), which can be related to energy pathways as CKB stores adenosine triphosphate (ATP) in creatine in neuronal cells and muscles, whereas valine can be converted into ATP in neuronal cells^43,48^. In mice, valine has been shown to inhibit CKB, providing orthologous evidence of a causal relationship^42^.

#### Regulation that has supporting evidence

Fifteen other enzyme-metabolite pairs have additional emerging evidence, either from animal studies, presence in related pathways, or there is reasoning on why the causal relationship may be valid, but no definitive human experimental result (**Supplementary Table 11**). Three combinations (PLA2G7 onto linoleate (18:2n6), linoleate (18:2n6) onto PLA2G7 and Glycine onto ornithine transcarboxylase (OTC)) were recurrently identified in 2 independent studies, representing independent lines of evidence for the causality of these relationships. Furthermore, we identified multiple bidirectional causal relationships between lipoprotein-associated phospholipase A2 (PLA2G7) and the essential fatty acid linoleate, as well as with cholesterol. MR-link-2 identified two linoleate to PLA2G7 causal relationships (PLA2G7 measured twice, two causal estimates: α̂ = 0.19, P = 9.7 . 10^-14^ and α̂ = 0.36, P = 1.3 . 10^-47^) and two causal relationships from the enzyme to the metabolite (PLA2G7 measured twice, two causal estimates: α̂ = 0.05, P = 4.7 . 10^-8^ and α̂ = 0.24, P = 2.2 . 10^-61^). There is emerging evidence linking PLA2G7 and linoleate: PLA2G7 releases linoleate from dorsal root ganglia ^49^ and PLA2G7 is protective of the peroxidation of conjugated linoleic acids ^50^. In line with this, we identify a causal relationship between cholesterol levels and PLA2G7, which has been associated with low-density lipoprotein particles, the primary cholesterol transporter in blood (Tellis and Tselepis, 2009)

#### Regulation without supporting evidence

For 62 pairs, our literature search did not find any supporting evidence for causal relationships. Some patterns suggest that they may be false positives, while others could reflect novel biology, which currently has limited evidence (**Supplementary Table 11**). One example involves eight links derived from two pQTL studies and two mQTL studies that link bilirubin and biliverdin to BCHE (range of α̂ = -0.1 to -0.15 , range of P = 2 . 10^-7^ to 9 . 10^-15^), which might represent confounding by liver damage, which causes both increases in BCHE and bilirubin^51^. Alternatively, there is weak evidence that bilirubin and biliverdin are related to choline esterase from in *vitro* experiments and in model organisms^52,53^.

### Integrating drugs and their effects into biological understanding

Using the OpenTargets resource, we matched drugs to their target proteins and assessed the downstream consequences of these proteins on metabolites^54^. Such an analysis provides insights into whether a drug has the intended effect on molecular phenotypes an can help identify if the perturbation of the gene that the drug targets has unintended side effects on metabolite levels^12^. Out of the 50 unique proteins that MR-link-2 identifies as having at least one causal effect, 11 are targets of compounds that are either registered as medicines (n = 20), are under investigation or have failed trials (n = 19) (**Supplementary Table 12**). One notable example is that MR-link-2 identifies a causal relationship between N-acetyl putrescine and IL1RAP (α̂ = 0.10, P = 2.7 . 10^-7^), which is targeted by the drug Spesolimab, used to treat psoriasis. Putrescine and other polyamines are present in psoriatic tissues, supporting the biological relevance of the estimated causal relationship^55,56^. It is worth noting that putrescine was not measured in the mQTL studies included in this work, and n-acetyl putrescine is one reaction away from putrescine (KEGG R01154).

## Discussion

In this study, we performed bidirectional MR between 213 metabolites and 506 enzymes to assess if MR is suitable to identify known biochemical reactions and discover putative novel enzymatic regulation of metabolites. In *cis* MR, all MR-methods perform similarly and have high precision (35-47%) and low recall (3.2%-4.6%), resulting in limited discriminative ability (AUC < 0.554). Extending our analyses to include genome-wide instrumental variables, we performed bidirectional MR between enzymes and metabolites. Discriminative ability remains low (AUC < 0.531), but we found a strong enrichment for direct pathway regulation among Bonferroni significant meta-analyzed MR results . We further focused on metabolite-protein links identified by MR-link-2, a method that we have developed which showed high precision in the Bonferroni significant results, and overall lower type 1 error rate and better recall/precision ratio compared to other methods^16^. We justify this focus as our aim was to understand how well MR methods can recover *bona fide* metabolite-enzyme relationships, rather than method comparison. Manual review of 106 Bonferroni significant enzyme-metabolite pairs identified 12 canonical reactions, eight reverse canonical reactions, four combinations with homologous evidence, six with pathway evidence and 23 with supporting evidence. For the remaining combinations we could not identify literature support. Finally, when matching 11 enzymes to therapeutics using OpenTargets, we identify the causal relationship between N-acetyl putrescine and IL1RAP that aligns with the therapeutic effect of the Spesolimab drug, used to treat psoriasis.

One important benefit of MR analysis is that the exposure and outcome do not need to be measured in the same individual^11^. This allows for a phenotype measured in one cohort to be tested for causality with a phenotype measured in another cohort. This provides the opportunity to study a much broader set of potential causal relations, such as bidirectional regulation between metabolites and enzymes or the metabolic downstream consequences of drugs. Still, we warrant caution in the interpretation of these analyses as MR results still suffer from large false positive rates and robust validation of regulatory pathway mechanisms would require targeted experimentation and case-control trials.

Indeed, there are substantial challenges to overcome before MR can be widely applied to study enzyme-metabolite links. An inherent limitation of large-scale metabolomics and proteomics studies coupled to the genotype information of the participants is that not all molecular species are measured. Even among those that are, not all are instrumentable in MR. A promising avenue is to use protein or metabolite ratios to understand intermediate pathway phenotypes^57,58^. These composite phenotypes are still understudied but could provide indications of unmeasured phenotypes or mechanisms in known pathways, even though analyzing ratio phenotypes comes with their own challenges^59^. A second major challenge is tissue specificity. Measurements underlying this study are derived from blood. Yet, most of human metabolism is tissue-specific, with certain tissues being more important for certain pathways^60^. It is still unclear how much our results will translate into other tissues, although the limited specificity of our analysis already provides evidence that we will not be able to recover most elements of pathway regulation in gold standard datasets.

There are still unknowns about how to conduct an appropriate MR analysis, for instance it’s unclear how much MR performance improvement can be expected from further increased sample size to derive molecular QTLs. Power analysis for MR studies usually do not consider locus selection mechanism and are hard to extend to pleiotropy robust methods^61,62^, making it difficult to distinguish true negatives from links where MR is simply underpowered.

As study sample size increase, it is likely that more causal links from other tissues can be proxied by blood, albeit with weaker effects. Here, it is important to consider that more subtle QTLs need to be analyzed with pleiotropy robust methods, as the mixture of different tissue effects can violate the exclusion restriction in MR. This study also highlights that the selection of suitable genetic regions for MR can be improved, as illustrated by the *cis* MR analysis, which has the highest precision and high recall, with the caveat that not all phenotype combinations can be tested. When extending to the *cis*+*trans* analysis, the recall (in terms of unique true positive findings) does not necessarily increase, even though some loci do provide correct inference. This indicates that putting biological priors on certain genomic regions may be the correct course of action to reduce type 1 error rate.

Taken together, our study indicates that there is strong potential for the analysis of molecular pathways using observational causal inference techniques, which is illustrated by our biologically informed findings. Such approaches are not only cost-effective, but their top-ranking metabolite-protein links are many-fold enriched for true associations, warranting experimental follow-up.

## Data Availability

The summary statistics used in this study are available from their respective publications (Methods). The genotype information underlying the LD matrices for the UK10K data resource were downloaded from the EGA under accession IDs EGAD00001000740 and EGAD00001000741. As this is individual level genotype data, a data access agreement is required for access. Conditions for this data access agreement can be found at https://www.uk10k.org/data_access.html. 

## Acknowledgements

We would like to thank all the study participants for their altruistic donations of their biological materials. Z.K. was funded by the Swiss National Science Foundation (SNSF 315230-219587).

## Author contributions

A.vd.G. and Z.K. conceptualized the study, A.vd.G. and S.R. performed data curation. A.vd.G. and Z.K performed formal analysis, including statistical, and computational techniques to synthesize and analyze data. Z.K. acquired funding. A.vd.G., S.R, C.A and Z.K. developed and designed methodology, Z.K. provided computational resources. A.vd.G. and S.R. performed software development. A.vd.G. created visualizations and wrote the original manuscript draft. A.vd.G., C.A., S.R and Z.K. reviewed, and edited the manuscript.

## Data and code availability

The summary statistics used in this study are available from their respective publications (**Methods**). The genotype information underlying the LD matrices for the UK10K data resource were downloaded from the EGA under accession IDs EGAD00001000740 and EGAD00001000741. As this is individual level genotype data, a data access agreement is required for access. Conditions for this data access agreement can be found at https://www.uk10k.org/data_access.html.

## Supplementary Figures

**Supplementary Figure 1:**
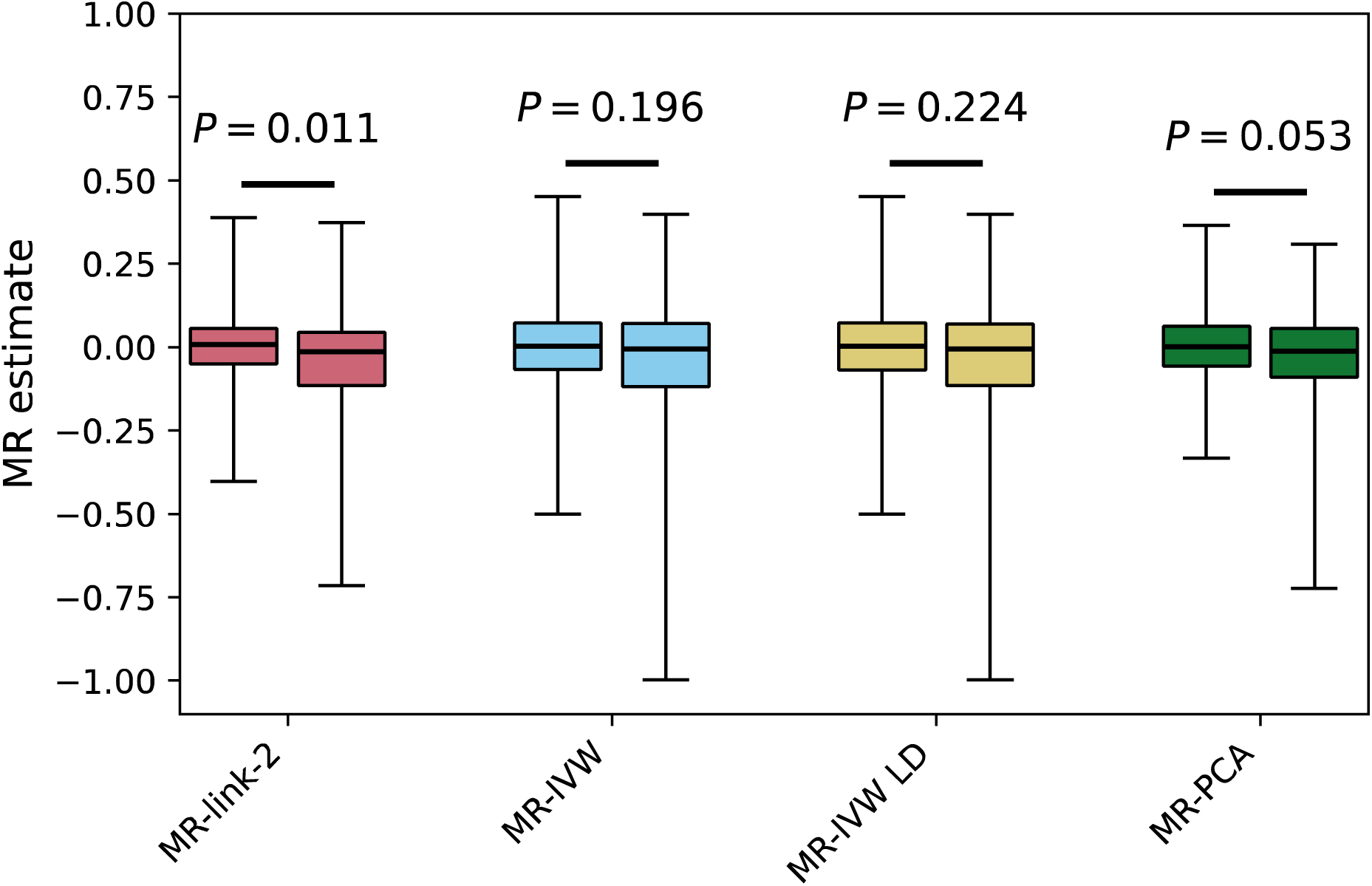
Causal estimates of enzymes onto products (left boxplot) and substrates (right boxplot) of for different MR methods. P values are based on a Kruskal-Wallis test. Middle line of the boxplots are medians, boxes contain 75% of all the datapoints, whiskers contain 95% of datapoints. Outliers outside these values are not shown.

**Supplementary Figure 2:**
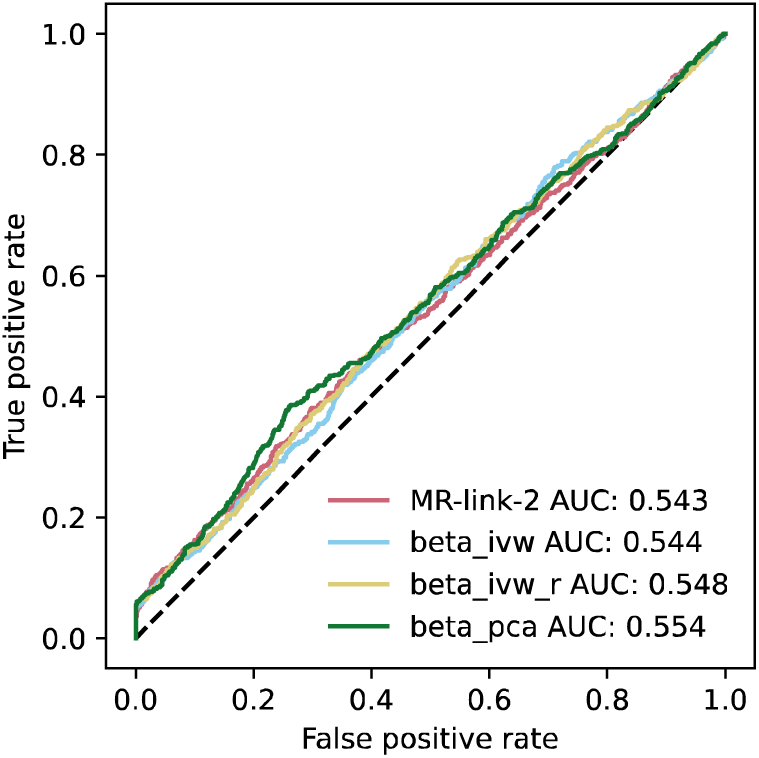
Discriminative ability of different MR methods from *cis* enzymes to their metabolites. (489 positives, compared to 25883 negatives).

**Supplementary Figure 3.**
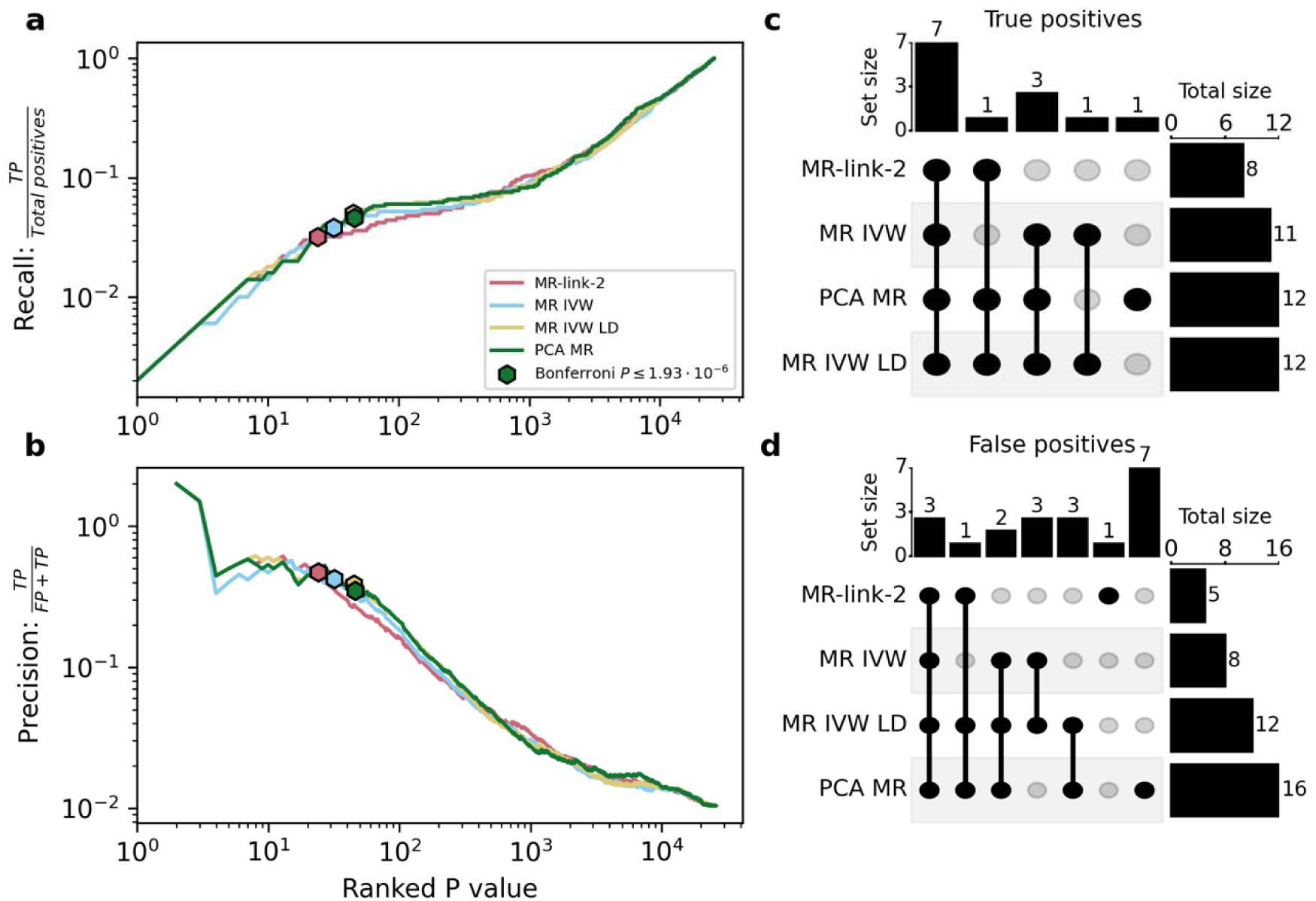
(**a**) recall and (**b**) precision curves of the cis MR methods tested in this study, only including genetic instruments in the cis regions of the focal protein, all valid comparison across studies are considered a single datapoint, using the region of the gene that transcribes the enzyme used in our metabolic map. (**c**) Set memberships for the true positives at each methods’ Bonferroni threshold. (**d**) Set memberships of false positives at each methods’ Bonferroni threshold. Abbreviations: (TP) True positive, (FP) False positive.

**Supplementary Figure 4:**
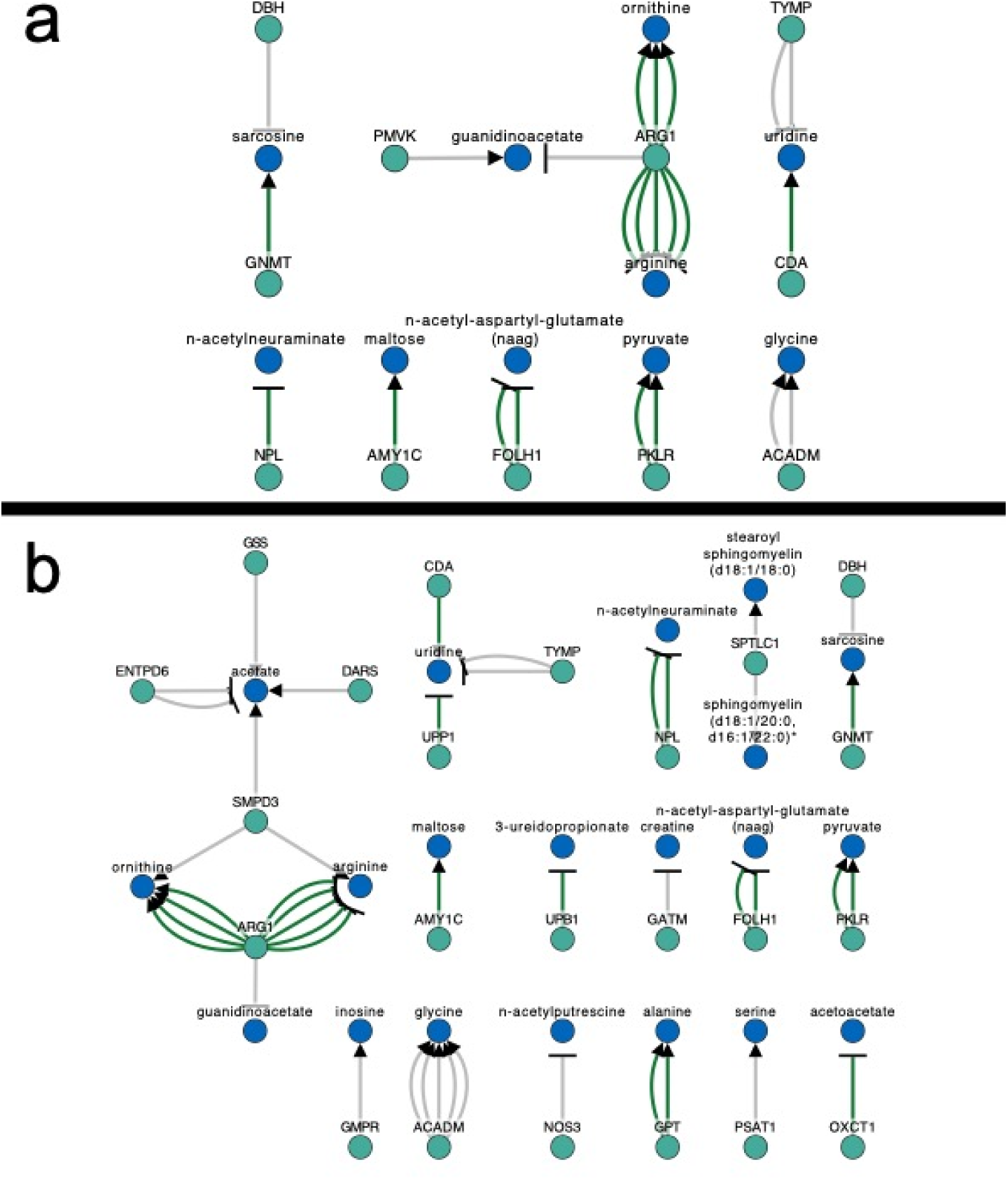
Graphs of significant *cis* MR results between enzymes and their reactants at Bonferroni significance. Panel a depicts the results for MR-link-2 and panel b depicts the results for MR-PCA. Depicting true positives (Green) from KEGG+Expasy and metacyc. False positives depicted in gray.

**Supplementary Figure 5.**
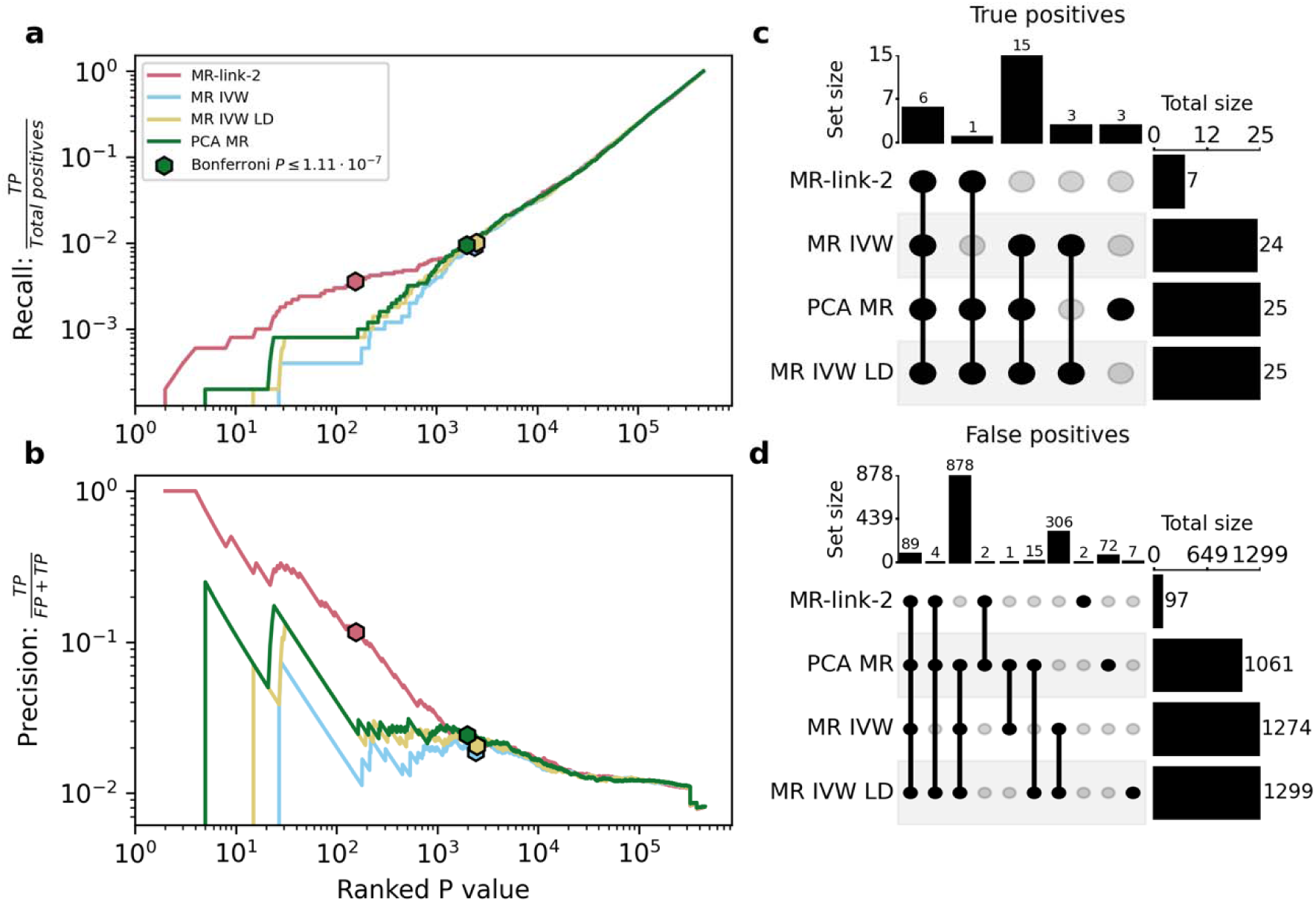
Precision and recall curves of MR methods when analyzing cis and non-cis regional results together in a bidirectional manner between enzymes and metabolites. (a) Recall and (b) precision depending on the chosen p value threshold ordering. (c) Set memberships for the true positives at each methods’ Bonferroni threshold. (d) Set memberships of false positives at each methods’ Bonferroni threshold .

## Supplementary Table legends

**Supplementary Table 1.** All retrieved reactions from both KEGG+expasy and Metacyc databases (database), with their respective reaction idenfier (reaction_id), enzyme commission enzyme number (enzymes) and their enzyme definition (definition).

**Supplementary Table 2.** All combined catalyzing proteins (Gene name, Ensembl ID, UNIPROT ID) measured in the two protein quantitative trait locus studies (study, accession) harmonized in this study, as well as where the enzyme comes from

**Supplementary Table 3.** All enzyme (Exposure accession, Exposure name) metabolite (Outcome accession, Outcome name) combinations that we can consider true positives in this study after harmonizing the ground truth datasets with the protein quantitative trait locus (QTL) and metabolite QTL studies, considering the enzyme (reaction reference, dataset) and if the metabolite is a product / substrate.

**Supplementary Table 4.** Cis-Mendelian randomization estimates per method (MR method), exposure (exposure accession, exposure name) and outcome (outcome accession) combination, with their respective MR estimate (alpha, se, p(alpha)), and meta-data on the origin of the exposure and outcome (pQTL dataset, uniprot ID, Metabolite dataset, metabolite HMDB ID), and finally if the combination is considered causal based on our ground truth dataset (causality).

**Supplementary Table 5.** Area under the receiver operator characteristic curve (AUC) for *cis*-Mendelian randomization estimates per MR-Method, combined with the number of positives in the datasets and the number of negatives

**Supplementary Table 6.** Brenda annotations (BRENDA Annotations) of Bonferroni significant (P < 1.93 . 10^-6^) (MR method) *cis* MR results (exposure, outcome, alpha, p(alpha)), their annotation in the ground truth dataset (causality), their metabolite and protein names (Metabolite name and Protein name)

**Supplementary Table 7.** All *cis+trans* regional Mendelian randomization results per method (MR method), associated region, exposure (exposure accession, Protein name) and outcome (outcome accession, Metabolite name) combination, with their respective MR estimate (alpha, se, p(alpha)), and meta-data on the origin of the exposure and outcome (protein dataset, UniProt ID, Metabolite dataset, metabolite HMDB ID, Ensembl ID), and finally if the combination is considered causal based on our ground truth dataset (causality).

**Supplementary Table 8.** All *meta-analyzed* combined Mendelian randomization results per method (MR method), associated region, exposure (exposure accession, Protein name) and outcome (outcome accession, Metabolite name) combination, with the number of meta-analyzed regions (n associated regions) and their respective meta-analyzed MR estimate (weighted alpha, weighted se, weighted p(alpha)), and meta-data on the origin of the exposure and outcome (protein dataset, UniProt ID, Metabolite dataset, metabolite HMDB ID, Ensembl ID), and if the combination is considered causal based on our ground truth dataset (causality).

**Supplementary Table 9.** Area under the receiver operator characteristic curve (AUC) for meta-analyzed Mendelian randomization estimates (MR) per MR-Method, combined with the number of positives in the datasets and the number of negatives

**Supplementary Table 10.** Enrichment results when comparing Bonferroni significant meta-analyzed results to all other results, with per method (MR method) the baseline probability of identifying a metabolic reactions (baseline probability), at Bonferroni significance (probability in Bonferroni significant), the resulting odds ratio and the fisher exact test P value

**Supplementary Table 11.** Bonferroni significant MR-link-2 results with all supporting evidence. Causal relationships are classified according to whether there is evidence of a canonical reaction, reverse canonical evidence (the metabolite influences the enzyme), pathway evidence (the metabolite and enzyme share a pathway), homologous reaction evidence (the enzyme has been shown to interact with the metabolite from homologous evidence) or Supporting evidence, when there is some evidence of a relationship existing, but it is not as strong as the other types. Enzymes or metabolites might be measured in multiple studies, leading to multiple estimates (indicated by a ‘2x’ or ‘3x’ in the “Causal relationship” column).

