## Supplementary Table 11 for "Mendelian randomization linking metabolites with enzymes reveals known and novel pathway regulation and therapeutic avenues"

***Supplementary Table 10** : 53 out of 106 Bonferroni significant MR-link-2 results with supporting evidence and interpretation classified according to whether there is evidence of a canonical reaction, reverse canonical evidence (where the metabolite influences the enzyme), pathway evidence (Where the metabolite and enzyme share a pathway), homologous reaction evidence (Where the enzyme has been shown to interact with the metabolite from homologous evidence) or suggestive evidence. Sometimes, the enzyme or metabolite is measured in more than one study leading to multiple estimates (indicated by a '2x' or '3x' in the Causal relationship column), we only report the most significant result and the number of Bonferroni signifcant ($P \leq 3.4 \cdot 10^{-7}$) combinations. MR-link-2 results are derived from **Supplementary Table 7***

| Evidence | Causal relationship (#significant estimates) | Causal estimate | P value | Interpretation |
|---|---|---|---|---|
| Canonical evidence | ARG1 → arginine (3x) | -0.66 | $6.8 \cdot 10^{-47}$ | Arginase 1 converts arginine into ornithine and urea (KEGG R00551) |
| Canonical evidence | ARG1 → ornithine (2x) | 0.39 | $1.4 \cdot 10^{-21}$ | Arginase 1 converts arginine into ornithine and urea (KEGG R00551) |
| Canonical evidence | NPL → N-acetylneuraminate | -0.37 | $2.3 \cdot 10^{-17}$ | N-acetylneuraminate pyruvate lyase converts N-acetylneuraminte into pyruvate and N-Acetyl-D-mannosamine (KEGG R01811) |
| Canonical evidence | PKLR → pyruvate (2x) | 0.21 | $1.7 \cdot 10^{-15}$ | Pyruvate kinase converts phosphoenolpyruvate into pyruvate and ATP (KEGG R00200) |
| Canonical evidence | AMY1(A\|B\|C)→ maltose | 0.17, | $2.8 \cdot 10^{-13}$ | The genes *AMY1A*, *AMY1B*, *AMY1C* encode for glycosylases that convert starch and related oligosacharides into maltose (KEGG R11262) |
| Canonical evidence | TYMP → 2'-deoxyuridine | -1.18 | $3.1 \cdot 10^{-18}$ | Thymidine phosphorylase converts deoxyuridine and orthophosphate into uracil and 2-deoxy-D-ribose 1-phosphate (KEGG R02484), missed in our reaction reference |
| Canonical evidence | ACY1 → N-acetylglutamate | -0.13 | $3.9 \cdot 10^{-8}$ | Aminoacylase converts N-acyl-amino acid (e.g., N-acetyl-glycine) into a carboxyl and amino acid (e.g., glycine). (KEGG R01263), missed in our reaction reference |
| Canonical evidence | ACY → glycine | -0.10 | $3.0 \cdot 10^{-7}$ | Aminoacylase converts N-acyl-amino acid (e.g., N-acetyl-glycine) into a carboxyl and amino acid (e.g., glycine). (KEGG R01263),missed in our reaction reference |
| Reverse canonical evidence | N-acetyl-aspartyl-glutamate → FOLH1 (2x) | -0.10 | $6.4 \cdot 10^{-40}$ | Glutamate carboxypeptidase II (FOLH1) converts N-acetyl-aspartyl-glutamate into N-acetyl-l-aspartyl-asparate and glutamate. (KEGG: R10687) |
| Reverse canonical evidence | Aspartate → DARS1 | 0.39 | $3.2 \cdot 10^{-7}$ | Aspartyl-tRNA synthetase (DARS1) converts tRNA and aspartate into aspartyl-tRNA. (KEGG R05577) |
| Reverse canonical evidence | Alanine → ACY1 | 0.30 | $3.2 \cdot 10^{-13}$ | Aminoacylase converts N-acyl-amino acid (e.g., N-acetyl-alanine) into a carboxyl and an amino acid (e.g., alanine) (KEGG R01263), missed In our reaction reference |
| Reverse canonical evidence | N-acetylneuraminate → NPL | -0.64 | $2.5 \cdot 10^{-8}$ | N-acetylneuraminate pyruvate lyase converts N-acetylneuraminte into pyruvate and N-acetyl-D-mannosamine (KEGG R01811) |
| Reverse canonical evidence | 2'-deoxyuridine → TYMP | -0.30 | $1.6 \cdot 10^{-7}$ | Thymidine phosphorylase converts deoxyuridine and orthophosphate into uracil and 2-deoxy-D-ribose 1-phosphate (KEGG R02484), missed In our reaction reference |
| Reverse canonical evidence | Arginine→ ARG1 (2x) | -0.32 | $1.2 \cdot 10^{-14}$ | Arginase 1 converts arginine into ornithine and urea (KEGG R00551) |
| Pathway evidence | TYMP → uridine (2x) | -0.57 | $3.3 \cdot 10^{-11}$ | Thymidine phosphorylase converts uracil into 2'-deoxyuridine. Uridine and 2'-deoxyuridine are separated by two reactions: uridine → uracil → 2'-deoxyuridine. |
| Pathway evidence | CYP3A4 → linoleate | 0.36 | $7.9 \cdot 10^{-21}$ | Cytochrome P450 3A4 is an oxidizing enzyme with a wide range of substrates, including fatty acids. Linoleate is a substrate of human recombinant CYP3A4 in human and rat liver microsomes. (Bylund et al., 1998) |
| Pathway evidence | Linoleate → CYP3A4 | 0.32 | $8.6 \cdot 10^{-32}$ | Cytochrome P450 3A4 is an oxidizing enzyme with a wide range of substrates, including fatty acids. Linoleate is a substrate of human recombinant CYP3A4 and human and rat liver microsomes. (Bylund et al., 1998) |
| Pathway evidence | 1-stearoyl-2-oleoyl-gps → PCYT2 | 0.57 | $9.8 \cdot 10^{-8}$ | Phosphate cytidylyltransferase 2, ethanolamine (PCYT2) is involved in phospholipid biosynthesis (Cikes et al., 2023). 1-stearoyl-2-oleoyl-gps (18:0/18:1) is a phospholipid intermediate, suggesting a feedback mechanism. |
| Pathway evidence | CKB → glycine | 0.17 | $8.2 \cdot 10^{-8}$ | Creatine kinase B phosphorylates creatine. Creatine is produced in a two-step reaction from glycine and L-arginine catalyzed by arginine:glycine amidinotransferase (AGAT) and guanidinoacetate methyltransferase (GAMT). (da Silva et al., 2009) |
| Homologous evidence | ALPI → Glucose | -0.03 | $2.6 \cdot 10^{-9}$ | Intestinal alkaline phosphatase (ALPI) is a wide specificity phosphoric-monoester hydrolase. Yeast orthologs can metabolize glucose. (Galabova et al., 2000) |
| Homologous evidence | ALPI → histidine | 0.04 | $2.9 \cdot 10^{-9}$ | Alkaline phosphatase intestinal (ALPI) is a wide specificity phosphoric-monoester hydrolase. Histidine was found to inhibit alkaline phosphatase in humans adipocytes. (Ali et al., 2006) |
| Homologous evidence | ALPI → Pyruvate | -0.03 | $2.0 \cdot 10^{-7}$ | Alkaline phosphatase intestinal (ALPI) is a wide specificity phosphoric-monoester hydrolase. Yeast orthologs can metabolize pyruvate. (Adler, 1978) |
| Homologous evidence | CKB → Valine | -0.17 | $1.9 \cdot 10^{-7}$ | In mice, valine inhibits creatine kinase B (CKB) activity (Pilla et al., 2003). Furthermore, CKB, which is primarly expressed in the brain, buffers ATP, while valine is a source of ATP in the brain (Kazak and Cohen, 2020), suggesting a feedback mechanism between the two sources of energy. |
| Supporting evidence | N-acetyl putrescine → IL1RAP | 0.10 | $2.7 \cdot 10^{-7}$ | IL1RAP is targeted by the drug Spesolimab, used to treat psoriasis. Putrescine is elevated in psoriasis and is separated from N-acetyl-putrescine by a single reaction (KEGG R01154) (LOWE et al., 1982; Morita et al., 2024; Ruseva et al., 2021). |
| Supporting evidence | N-acetylneuraminate → ALPI | 0.16 | $1.0 \cdot 10^{-7}$ | N-acetylneuraminate is a major component of the intestinal mucus layer, while intestinal alkaline phosphatase (ALPI) helps maintains gut barrier function(Bell et al., 2019; Fawley and Gourlay, 2016). |
| Supporting evidence | PCYT2 → pyruvate | 0.23 | $9.1 \cdot 10^{-8}$ | Phosphate cytidylyltransferase 2, ethanolamine (PCYT2) converts cytidine triphosphate and ethanolamine phosphate into diphosphate and cytidine diphosphate ethanolamine in the Kennedy pathway of phosphatidylethanolamine synthesis. Older *Pcyt2*$^{+/-}$ mice have increased rates of liver gluconeogenesis, which converts pyruvate into glucose (Grapentine et al., 2022) |
| Supporting evidence | GAPDH→ sphingosine | 2.00 | $3.4 \cdot 10^{-7}$ | Glyceraldehyde-3-phosphate dehydrogenase (GAPDH) is a rate limiting enzyme in glycolysis . In mice knockouts that cause increased sphingosine, glycolysis is upregulated. In humans, sphingosine-1-phosphate |

| | | | | |
|---|---|---|---|---|
| | | | | increases glycolysis in hypoxic circumstances (Sun et al., 2016). Suggesting a feedback mechanism between sphingosine and GAPDH (Sun et al., 2017). |
| Supporting evidence | PLA2G7 → linoleate (18:2n6) (2x) | 0.24 | $2.3 \cdot 10^{-61}$ | Phospholipase A2 group VII (PLA2G7) and linoleate are related to another: PLA2G7 releases linoleate from dorsal root ganglia (Boyd et al., 2021) and PLA2G7 is protective of the peroxidation of conjugated linoleic acids (Vermonden et al., 2024). |
| Supporting evidence | PLA2G7 → cholesterol | 0.30 | $9.2 \cdot 10^{-10}$ | Phospholipase A2 group VII (PLA2G7) is associated with low-density lipoprotein particle, the primary cholesterol transporter in blood (Tellis and Tselepis, 2009) |
| Supporting evidence | linoleate (18:2n6) → PLA2G7 (2x) | 0.36 | $1.4 \cdot 10^{-42}$ | Phospholipase A2 group VII (PLA2G7) and linoleate are related to another: PLA2G7 releases linoleate from dorsal root ganglia (Boyd et al., 2021) and PLA2G7 is protective of the peroxidation of conjugated linoleic acids (Vermonden et al., 2024). |
| Supporting evidence | DBH → sarcosine | -0.06 | $2.2 \cdot 10^{-9}$ | Dopamine beta-hydrolase (DBH) converts dopamine into norepinephrine, both of which act as neurotransmitters. Dopamine levels are relevant to schizophrenia (Brisch et al., 2014; Kesby et al., 2018). Sarcosine acts as an adjuvant to many schizophrenia treatments (Lane et al., 2006; Pawlak et al., 2023; Tsai et al., 2004). |
| Supporting evidence | flavin adenine dinucleotide → PCYOX1 | 2.35 | $2.0 \cdot 10^{-20}$ | Prenylcysteine oxidase (PCYOX1) is a flavoprotein that contains the flavin adenine dinucleotide as a cofactor (Tschantz et al., 2001). |
| Supporting evidence | RAB5B → sphingosine | 1.60 | $1.7 \cdot 10^{-7}$ | Ras related protein 5B (RAB5B) is a GTPase is an enzyme involved in endocytosis in conjunction with other RAB5B isoforms(Bucci et al., 1995). Evidence is emerging that Shingosine is also involved in endocytosis: sphingosine kinase 1 (SPHK1) colocalizes with RAB5 proteins on endosomes, and sphingosine is a substrate for SPHK1 (Shen et al., 2014). 7/29/25 5:00:00 PM |
| Supporting evidence | Glycine → OTC (2x) | 0.07 | $6.6 \cdot 10^{-10}$ | Ornithine transcarbamylase (OTC) is an enzyme of the urea cycle converting ornithine and carbamoyl phosphate into citrulline and phosphate. The glycine cleavage system represents one of the key glycine catabolic pathways and generates ammonia (Wang et al., 2013). The latter is metabolized and excrete by the urea cycle. |
| Supporting evidence | Glycine → FBP1 | 0.07 | $1.6 \cdot 10^{-10}$ | Glycine is a gluconeogenic amino acid, while fructose-1,6-bisphosphatase 1 (FBP1) is a gluconeogenic enzyme (D'Andrea, 2000; Visinoni et al., 2012). |
